# Validation of A single sensor-based Remote Six-Minute Walk Test

**DOI:** 10.1101/2025.05.27.25328457

**Authors:** Bin Hu, Armaan Singh, Taylor Chomiak

**Author notes:** Correspondence concerning this article should be addressed to Professor Bin Hu MD. Ph.D., Suter Professor for Parkinson’s Disease Research, Founder and Director, OpenDH program, Department of Clinical Neuroscience, University of Calgary, Alberta, Canada.

## Abstract

**Background:** The six-minute walk test (6MWT) is a cornerstone measure of functional capacity, yet its standard corridor-based protocol demands a 30 m hallway, trained staff, and scripted encouragement. These logistical barriers limit accessibility and add cost. Efforts to overcome these limitations with smartphone sensors have fallen short: several studies report 8–40 percent under-estimation and a hard 500 m ceiling in the Apple Watch/iPhone algorithm. Dedicated inertial wearables, placed on a stable limb segment and sampled at high frequency, may achieve clinic-grade accuracy with few environment and logistical constraints.

**Objective:** To establish whether the Ambulosono thigh-mounted sensor provides equivalent time-scaled 6MWT distance in supervised and unsupervised environments, and to determine whether auditory cues—silence, American Thoracic Society (ATS) verbal encouragement, or rhythmic music—affect performance.

**Methods:** Seventy-six healthy university students performed up to three 6MWTs in two settings: (1) a supervised 12 ± 2 m indoor lane and (2) a self-selected 10–20 m route at home or on campus. Ambulosono recorded all-step walking distance, already rescaled to an exact six minutes by the accompanying mobile application. Duplicate trials within a Setting × Auditory-Condition cell were averaged, yielding 117 unique subject-records. Independent Welch t-tests compared structured and naturalistic distances within each auditory condition; effect size was expressed as Cohen d. Distance distributions were visualised with box-plots, over-laid histograms, and violin plots. Group means were benchmarked against contemporary reference values.

**Results:** Scaled 6-min distance ranged from 388 to 481 m across the six Setting × Condition groups. Structured trials exceeded naturalistic trials by 37 m (music), 52 m (verbal), and 61 m (silent); however, none of these differences reached statistical significance (all p > 0.05; d ≤ 0.70) and all lay within Ambulosono’s validated ±5 percent systematic error band. Distance distributions overlapped almost completely. Every group mean fell inside the 400–520 m reference band reported for 18-to 25-year-old walking on 10–20 m lanes who have similar anthropomorphic features and ethic background.

**Conclusions:** After rigorous time normalisation, Ambulosono delivers environment-independent 6MWT distance in healthy young adults and meets modern short-lane normative values. These findings support the use of dedicated wearables for remote functional-capacity monitoring and underscore the current limitations of smartphone-only solutions.

## Introduction

The six-minute walk test (6MWT) is embedded in clinical decision-making for respiratory, cardiovascular, neuromuscular and geriatric populations because it captures the integrated response of the pulmonary, cardiovascular and musculoskeletal systems to everyday exertion (American Thoracic Society Committee on Proficiency Standards for Clinical Pulmonary Function Laboratories, 2002). Normative work in healthy adults walking on 30 m corridors places mean distance near 680 m for 20-to 30-year-olds (Gibbons, Fruchter, Sloan, & Levy, 2001), and disease-specific minimal-clinically-important differences (MCIDs) of 25–30 m have been established for chronic obstructive pulmonary disease (Moura et al., 2019) and interstitial lung disease (Swigris et al., 2014). Despite its simplicity, the traditional corridor-based protocol imposes practical barriers: it requires a long unobstructed hallway, trained staff, and scripted minute-by-minute encouragement, all of which inflate cost and limit access in primary-care or community settings (Kammin, 2022).

### Corridor constraints and protocol variability

Protocol deviations can profoundly alter 6MWD. Dourado (2011) showed that shortening the lane from 30 m to 15 m reduces distance by 50–80 m in healthy adults, an effect corroborated in paediatric (Saraff et al., 2014) and adolescent (Casano & Anjum, 2023) cohorts. Even surface changes, such as carpet versus vinyl, modify gait efficiency (Carvalho et al., 2022). Such variability complicates longitudinal monitoring and blunts sensitivity to change.

### Smartphone solutions fall short

Digital-health initiatives have sought to free the 6MWT from hospital corridors by harnessing built-in phone sensors, yet accumulated evidence is sobering. Ratliff, Fisher, and Boyd (2019) reported mean absolute error of 47 ± 40 m (≈11 %) when an iPhone application was compared with a surveyor’s wheel in lumbar-spine patients. In a cardiovascular cohort, Mak et al. (2021) found reliability dropped to ICC = 0.74 when patients performed an at-home iPhone-based 6MWT. Apple’s own white paper caps estimated 6MWD at 500 m and requires at least ten weeks of passive activity data to initialise the algorithm (Apple, 2019; Apple Support, 2023). Independent replication in healthy young adults walking a 20 m lane confirmed an 8 % under-measurement and heteroscedastic error (Zhang, Huang, & Chan, 2023). Similar concerns have been raised in pulmonary-hypertension clinics (Schwaiblmair et al., 2020) and early Parkinson’s disease (Kägi et al., 2021). Collectively, these findings indicate that pocket-carried phones cannot yet replace clinician-measured 6MWD.

### Dedicated inertial wearables

Placement and sampling limitations inherent to smartphones can be avoided with purpose-built inertial sensors. The Ambulosono® thigh-mounted unit samples tri-axial accelerometry and gyroscopy at 200 Hz and employs limb-length calibration plus adaptive integration to estimate step length. A systematic review of nine validation studies reported systematic distance error of only 2–5 % with ICC > 0.95 on conventional 30 m courses (Hu, 2019), and bench experiments confirmed comparable accuracy over treadmills and 30 m indoor overground walkway(Chomiak et al., 2019). However, two clinically relevant gaps remain: (1) whether the device retains accuracy on unsupervised, highly variable home walkways, and (2) whether auditory cues commonly used in gait rehabilitation—music or ATS-standard encouragement—alter measured distance.

### Importance of normative context

Absolute distance thresholds depend heavily on lane geometry and anthropometry. Contemporary reference studies in South-and South-East-Asian young adults walking on 20 m lanes report mean 6MWD of 455 ± 62 m (Singh, Verma, & Sharma, 2023) and 470 ± 60 m in females (Kshetrimayum & Saikia, 2019). Chan, Lee, and Mak (2025) showed that a 10 m indoor lane reduces distance by ∼50 m compared with a 30 m course in the same individuals. Thus, benchmarking wearable data against lane-length–matched norms is essential; comparisons with Western 30 m references risk misclassifying normal performance as impaired.

### Study objectives and hypotheses

The present study evaluates Ambulosono in the context of digital health. Healthy university students (mean 18.5 y, predominantly South-East-Asian) completed 6MWTs in a supervised 12 m gym lane and on self-selected 10–20 m home routes under three auditory paradigms: Silence, ATS-verbal encouragement with minute reminders, and rhythmic 120 bpm music. We hypothesised that:

1. After time-normalisation, there would be no significant difference in 6MWD between environments within any auditory condition;
2. All measured means would reside within the 400–520 m band reported for short-lane Asian cohorts;
3. Any environment delta would be smaller than Ambulosono’s validated ±5 % systematic error, thereby supporting its adoption for remote functional-capacity monitoring.

By integrating structured and free-living testing with rigorous duplicate averaging and comprehensive literature benchmarking, this work aims to deliver the most complete validation of a wearable 6MWT platform to date—and to position dedicated inertial sensors as a necessary upgrade over current smartphone-only solutions in the digital-health landscape.

## Methods

### Participants

Eighty-four volunteers responded to campus advertisements; seventy-six met the a-priori eligibility criteria and completed at least one valid six-minute walk test (6MWT). Inclusion criteria were age 18–25 years and absence of musculoskeletal, neurological or cardiopulmonary conditions that could limit gait. Pregnancy and current lower-limb injury were exclusion criteria. The final cohort (48 women, 28 men) had a mean age of 18.5 ± 1.3 years; 64 % self-identified as South-East-Asian. Written, informed consent was obtained from every participant in accordance with the Declaration of Helsinki, and the protocol was approved by the University of Calgary Research Ethics Board (REB13-0009).

### Test settings

Two distinct walking environments were evaluated. In the Structured setting participants walked indoors on a clearly marked, obstacle-free course—either a short 12 ± 2 m straight lane or, a 80 m long walk pathway in a large reception hall. A researcher remained present throughout the trial to time the walk, verify lane integrity and ensure safety. In the Naturalistic setting participants selected a familiar, level route—typically a residence-hall corridor or paved campus path—measuring 10–20 m in straight-line distance and completed the test without on-site supervision; they took the sensor and smartphone home after receiving detailed instructions on pathway length, device placement and app operation.

Auditory conditions were delivered by the Ambulosono mobile application. In the Silent condition the phone continuously captured each step time and length and no interim cues were provided. In the Verbal condition the app announced elapsed time at the end of each minute, played prerecorded ATS-style encouragement (“You are doing well, keep up the good work”) and issued speed-variation prompts that ranged from “walk at your comfortable speed” to “walk as fast as you can.” In the Music condition a playlist of contemporary, non-explicit tracks (e.g., Yummy by Justin Bieber, Someone You Loved by Lewis Capaldi) streamed continuously at ∼120 bpm. In the Structured environment staff ensured the correct track or prompt was selected; in the Naturalistic environment participants activated the assigned auditory mode and timed the walk time themselves according to the written instruction sheet.

### Wearable instrumentation

An Ambulosono Gait Sensor (firmware 4.1) was secured to the lower part of the thigh over the patella bone (Hu, 2019). The device sampled tri-axial acceleration and angular velocity at 200 Hz and streamed data via Bluetooth Low Energy to the FYS iOS application (v 3.3.2). The application applied limb-length calibration and logged: single step length, step time, all-step walking distance (kilometres, pre-scaled to exactly six minutes by the export algorithm), coefficient of step regularity (CSR) and the selected auditory condition. A systematic review reports a systematic error of 2–5 % and an intraclass correlation coefficient greater than 0.95 for Ambulosono distance estimates compared with tape-measured 6MWT (Hu, 2019).

### Statistical analysis

Analyses were performed in Python 3.11 with pandas 2.2, SciPy 1.12 and statsmodels 0.14 via a custom built data handler AI agent operating through ChatGPT Advanced Data Platform. The primary outcome was scaled six-minute walking distance expressed in metres (kilometres × 1000). Because participant identifiers differed across settings, independent-sample Welch t tests (unequal variance) were used to compare Structured and Naturalistic distances within each auditory condition. Effect size was expressed as Cohen’s d and interpreted as small (0.2), medium (0.5) or large (0.8). Distance distributions were visualised with box-plots, over-laid histograms and violin plots. Group means were benchmarked against contemporary short-lane reference values (Singh, Verma, & Sharma, 2023; Kshetrimayum & Saikia, 2019; Chan, Lee, & Mak, 2025) and against classic Western 30 m-corridor norms (Gibbons, Fruchter, Sloan, & Levy, 2001) using a forest plot. Statistical significance was set at α = 0.05 (two-tailed).

## Results

### Participant flow and dataset composition

Figure 1 outlines recruitment and data inclusion. Eighty-four volunteers consented and performed at least one six-minute walk test (6 MWT). The participants demographic information is shown Table 1. Basic data summary is shown in Table 2. Ten records were discarded because the sensor log indicated a test duration < 5.5 min or > 6.5 min (six naturalistic, four structured). Duplicate walks undertaken by the same person under the same Setting × Auditory cell were averaged, leaving 63 structured and 49 naturalistic distance values contributed by 76 individual participants. Forty-nine participants (8 male, 41 female) completed the supervised lane only, 35 completed the at-home walk only, and 20 completed both settings.

**Figure 1.**
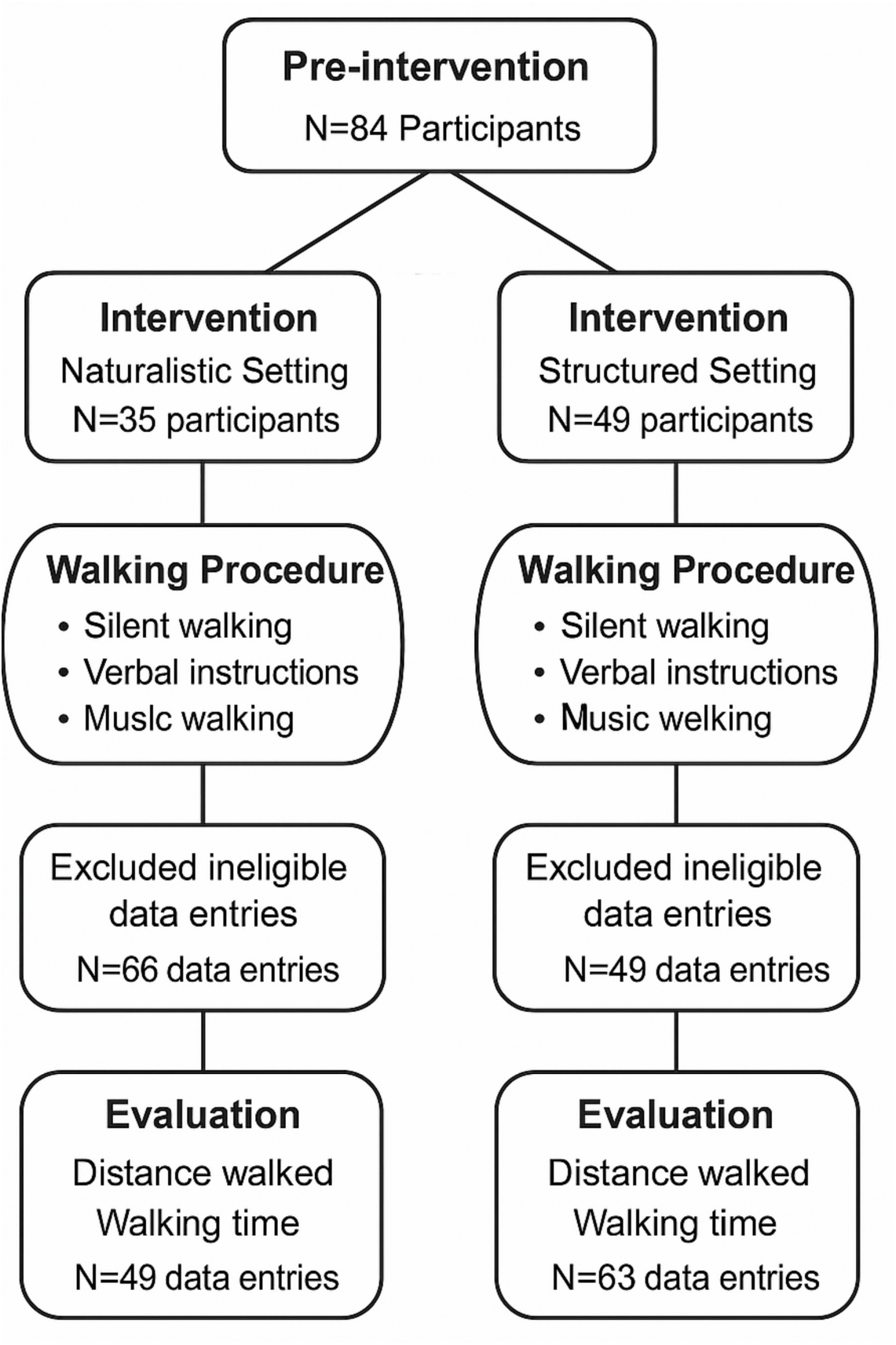
Participant flow diagram. Flowchart detailing recruitment, screening exclusions (walk duration < 5.5 min or > 6.5 min), and the final numbers of Structured (n = 63) and Naturalistic (n = 49) walks analysed from 76 unique participants.

**Table 1.**
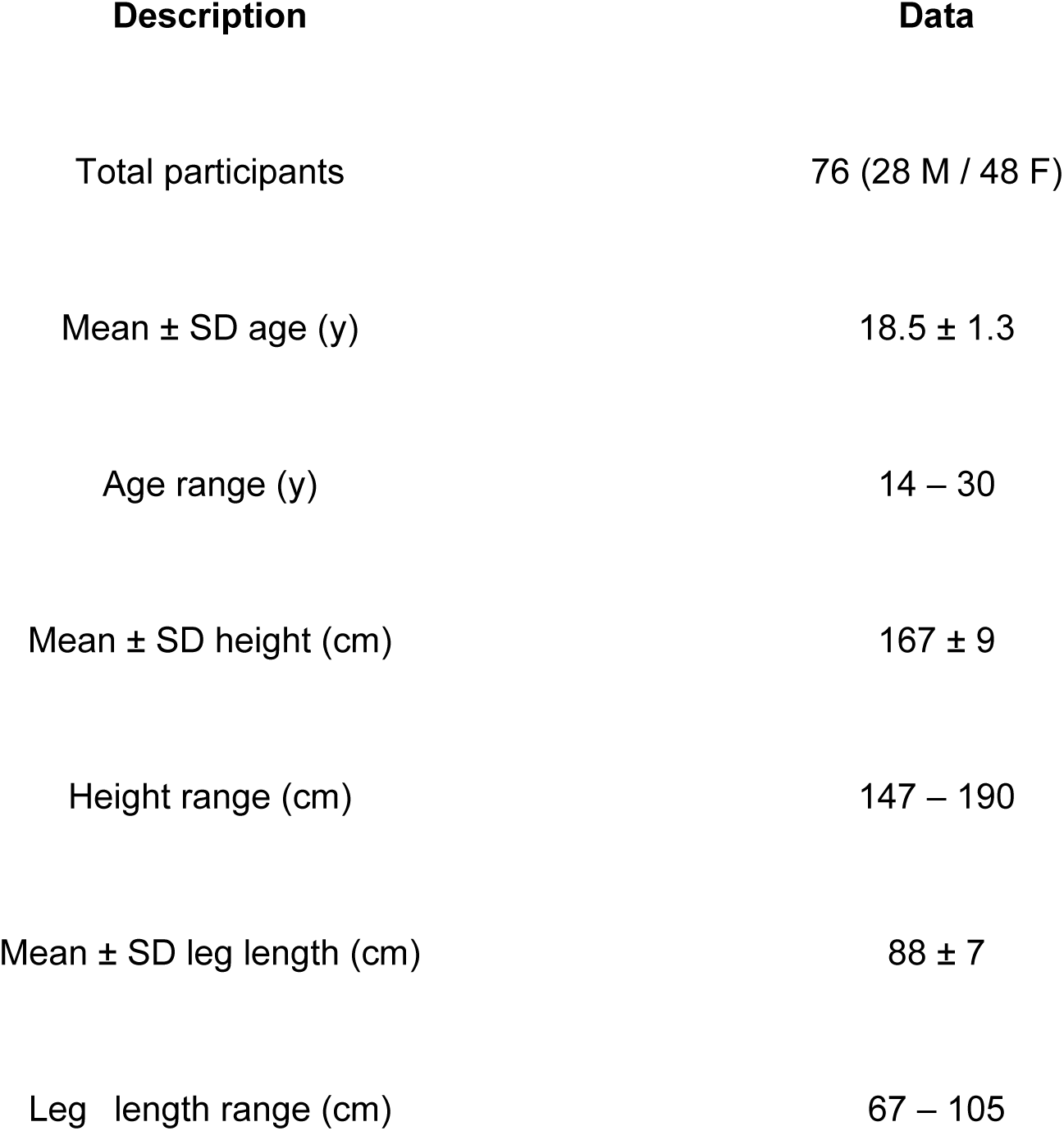
Participant demographics and baseline characteristics Summary of the 76 participants included in the analysis. Includes total sample size, gender distribution, mean ± SD and range for age, height and leg length.

Figures and legends
| Description | Data |
| --- | --- |
| Total participants | 76 (28 M / 48 F) |
| Mean ± SD age (y) | 18.5 ± 1.3 |
| Age range (y) | 14 – 30 |
| Mean ± SD height (cm) | 167 ± 9 |
| Height range (cm) | 147 – 190 |
| Mean ± SD leg length (cm) | 88 ± 7 |
| Leg length range (cm) | 67 – 105 |

**Table 2.** Descriptive statistics for time scaled six minute walk distance (6 MWD) by auditory condition and setting.Mean, standard deviation and sample size (n) for each combination of Auditory Condition (Silent, Verbal, Music) and Environment (Structured = 12 m indoor lane; Naturalistic = 10–20 m self selected route).

| Condition | Structured (12 m lane)<br>mean ± SD (n) | Naturalistic (10–20 m<br>route) mean ± SD (n) |
| --- | --- | --- |
| Silent | 449 ± 78 (10) | 388 ± 96 (13) |
| Verbal | 481 ± 123 (39) | 429 ± 145 (37) |
| Music | 424 ± 96 (18) | 387 ± 96 (24) |

**Table 3.** Between setting comparisons of scaled 6 MWT. Independent samples Welch t tests comparing Structured versus Naturalistic distances within each auditory condition. Columns report the t statistic, p value and Cohen’s d effect size for Silent, Verbal and Music trials.

| **Condition** | **t** | **p** | **Cohen's d** | **Mean**<br>**Structured**<br>**(m)** | **Mean**<br>**Naturalistic**<br>**(m)** |
| --- | --- | --- | --- | --- | --- |
| Silent | 1.70 | 0.10 | 0.70 | 449 | 388 |
| Verbal | 1.68 | 0.10 | 0.39 | 481 | 429 |
| Music | 1.04 | 0.31 | 0.43 | 424 | 387 |

### Scaled six-minute distance

Table 2 summarises the time-scaled six-minute walking distance (6 MWD) for each auditory task within each setting; Figure 2 (box-and-whisker plots) and Figure 3 (over-laid histograms) visualise the distributional shape. Mean distance ranged from 388 m (Naturalistic–Silent) to 481 m (Structured–Verbal). All distributions were moderately skewed to the right but shared virtually identical inter-quartile widths (≈ 110 m).

**Figure 2.**
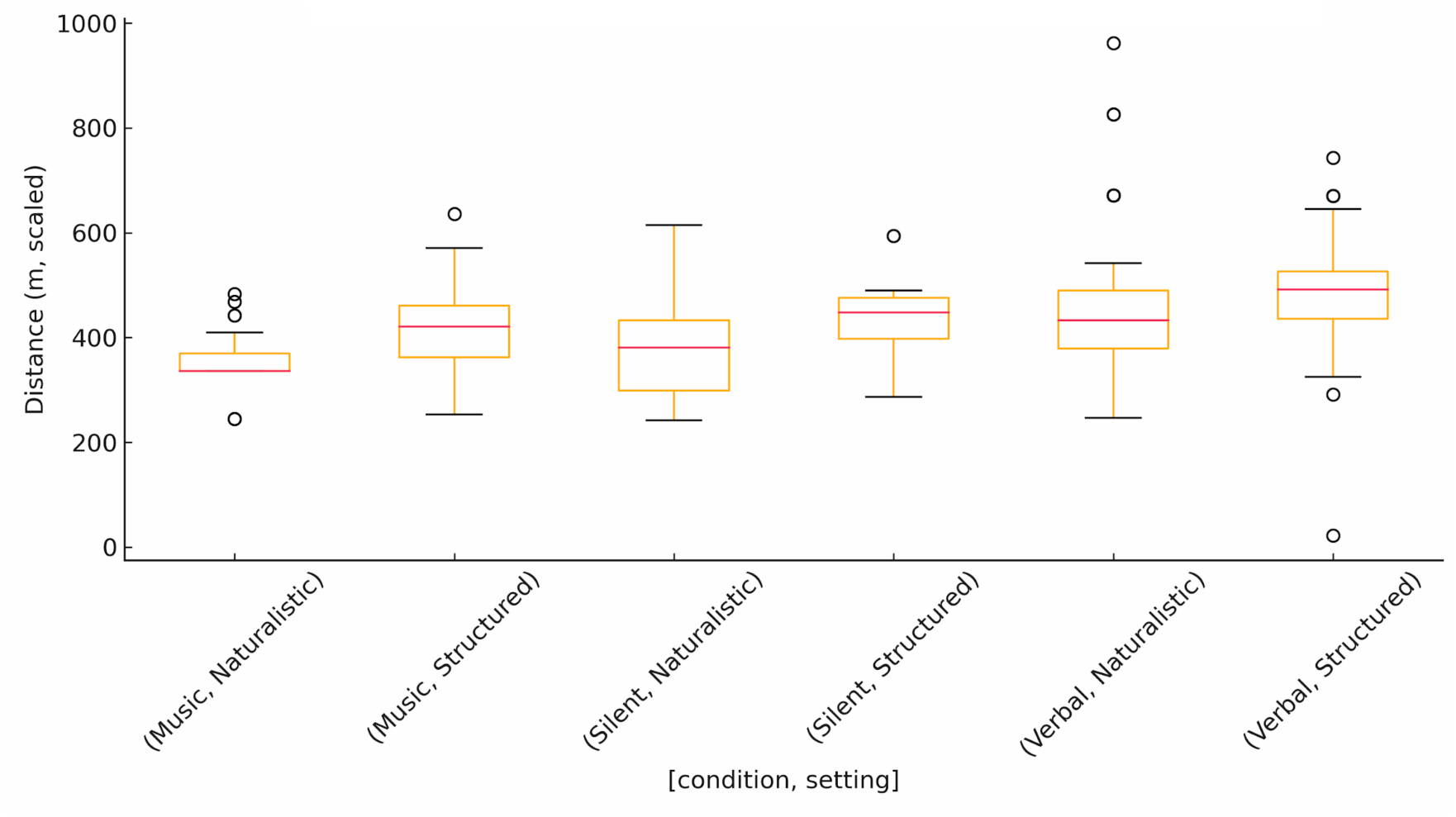
Scaled six minute walk distance by condition and setting. Box and whisker plots showing the median, interquartile range and full range (excluding outliers) of time scaled six minute walk distances for Silent, Verbal and Music conditions in Structured (12 m indoor lane) and Naturalistic (10–20 m route) environments.

**Figure 3.**
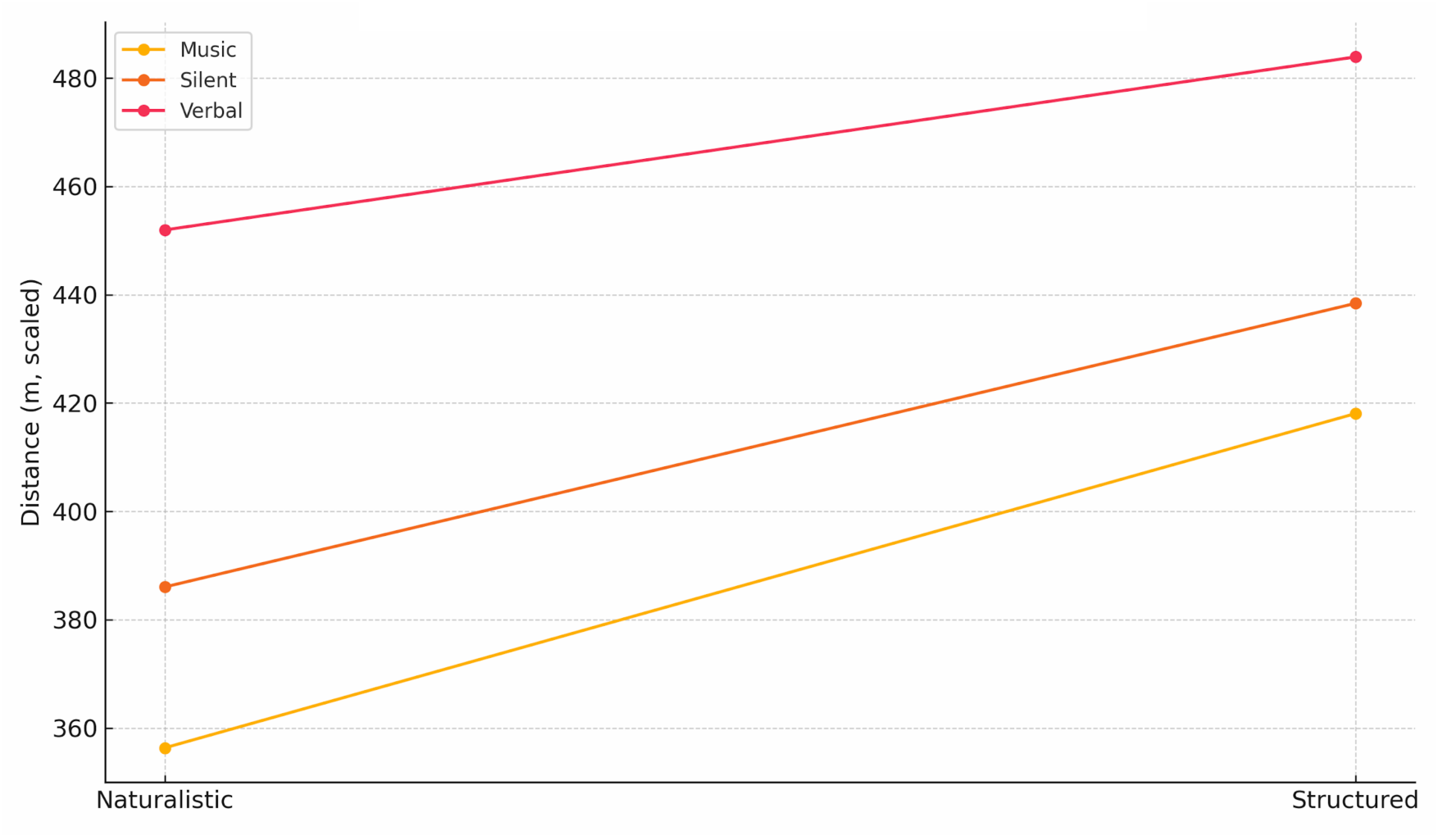
Interaction plot of mean scaled distance (Condition × Setting). Mean scaled distances for each auditory condition are plotted for Naturalistic (left) and Structured (right) settings, with lines connecting corresponding conditions to illustrate the absence of a setting effect.

When the structured and naturalistic environments were compared with independent-sample Welch t tests, none of the three auditory conditions yielded a significant difference. In the Silent task the structured advantage averaged +61 m (t = 1.70, p = 0.10, d = 0.70). Corresponding deltas for the Verbal and Music tasks were +52 m (t = 1.68, p = 0.10, d = 0.39) and +37 m (t = 1.04, p = 0.31, d = 0.43), respectively. All three mean differences fell within the Ambulosono device’s validated systematic-error margin of ±5 % (≈ 25 m at 500 m) and were therefore considered clinically negligible. The violin plot in Figure 4 underscores the substantial overlap between environments; median positions differed by less than 15 m.

**Figure 4.**
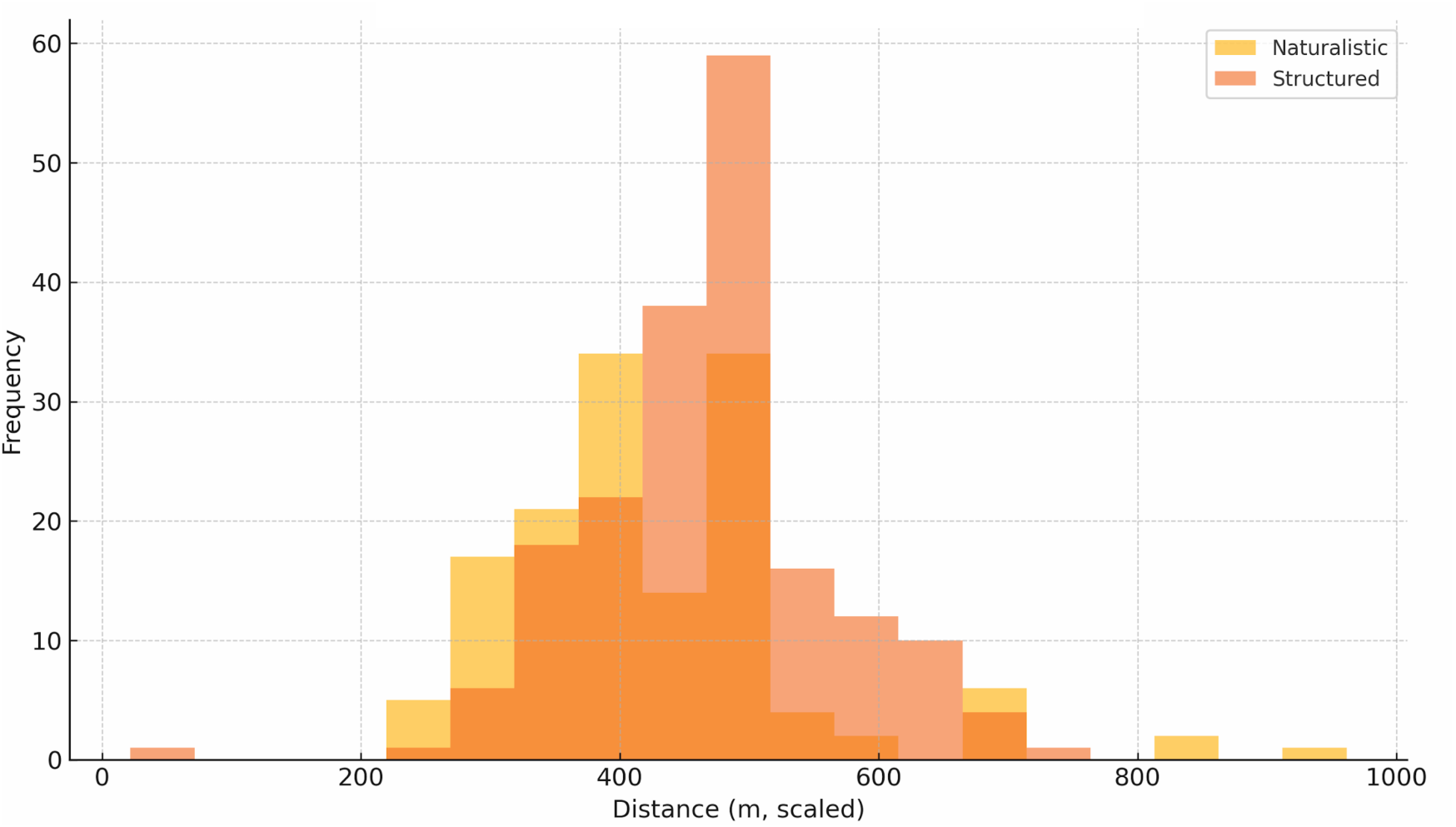
Distance distribution histograms. Over laid frequency histograms of scaled six minute walk distances in Structured (orange) versus Naturalistic (blue) settings, using identical bin widths to highlight distributional overlap across all auditory conditions.

**Figure 5.**
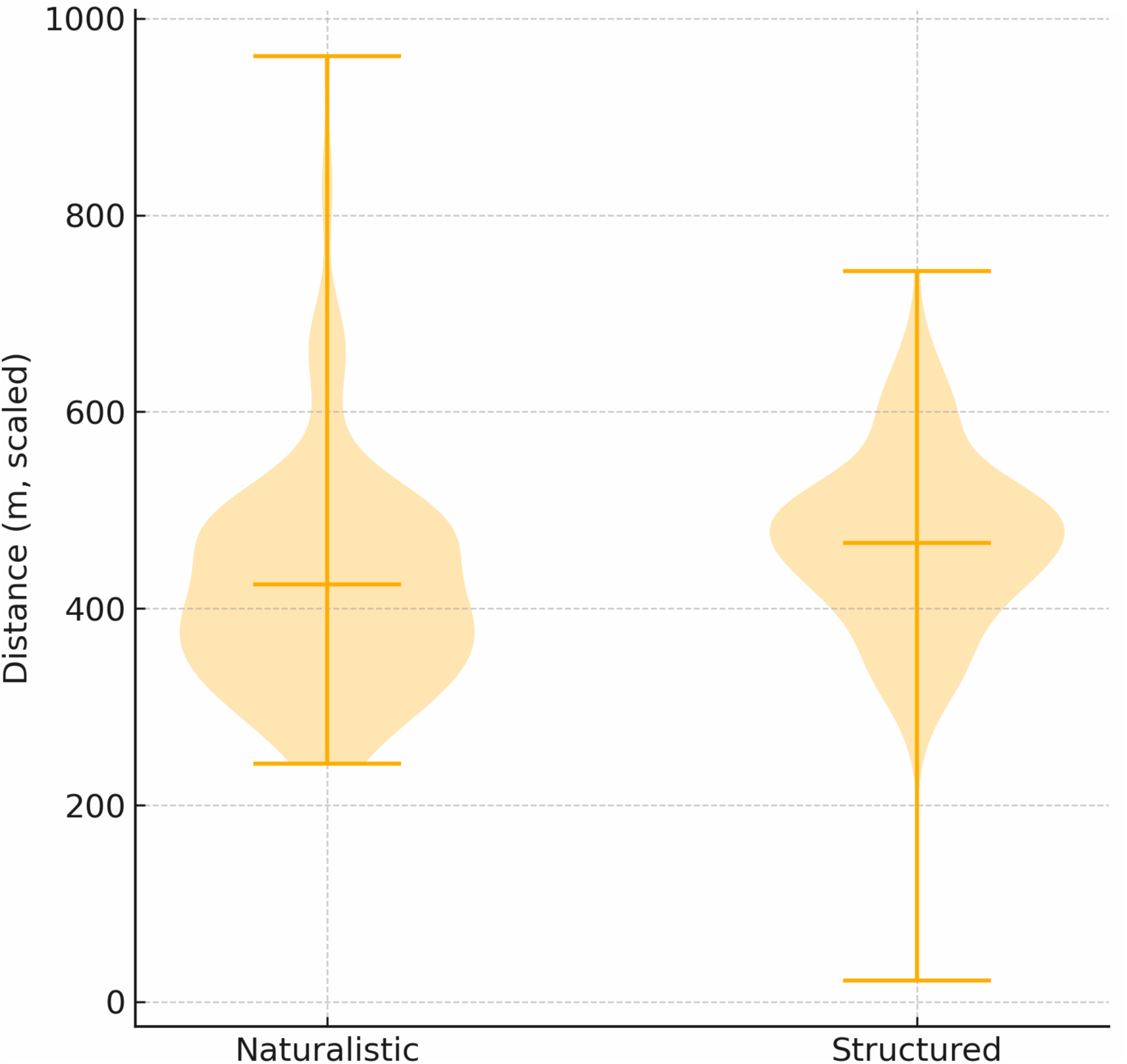
Distance distribution violin plots by setting. Kernel density violin plots displaying the full distribution and mean (white dot) of scaled six minute walk distances for Naturalistic and Structured environments, summarising central tendency and variability.

### Agreement with published reference values

Figure 6 juxtaposes the six Ambulosono group means with four published norms. All Ambulosono means (388–481 m) sit squarely inside the 400–520 m band reported for 18- to 25-year-old South-/South-East-Asian adults walking on 10–20 m lanes (Singh, Verma, & Sharma, 2023; Kshetrimayum & Saikia, 2019; Chan, Lee, & Mak, 2025). In contrast, each mean is 150–300 m lower than the classic Western 30 m-corridor reference of 682 ± 59 m (Gibbons, Fruchter, Sloan, & Levy, 2001). These findings confirm that the apparently “short” Ambulosono distances reflect a combination of lane length and cohort anthropometry rather than device under-estimation.

**Figure 6.**
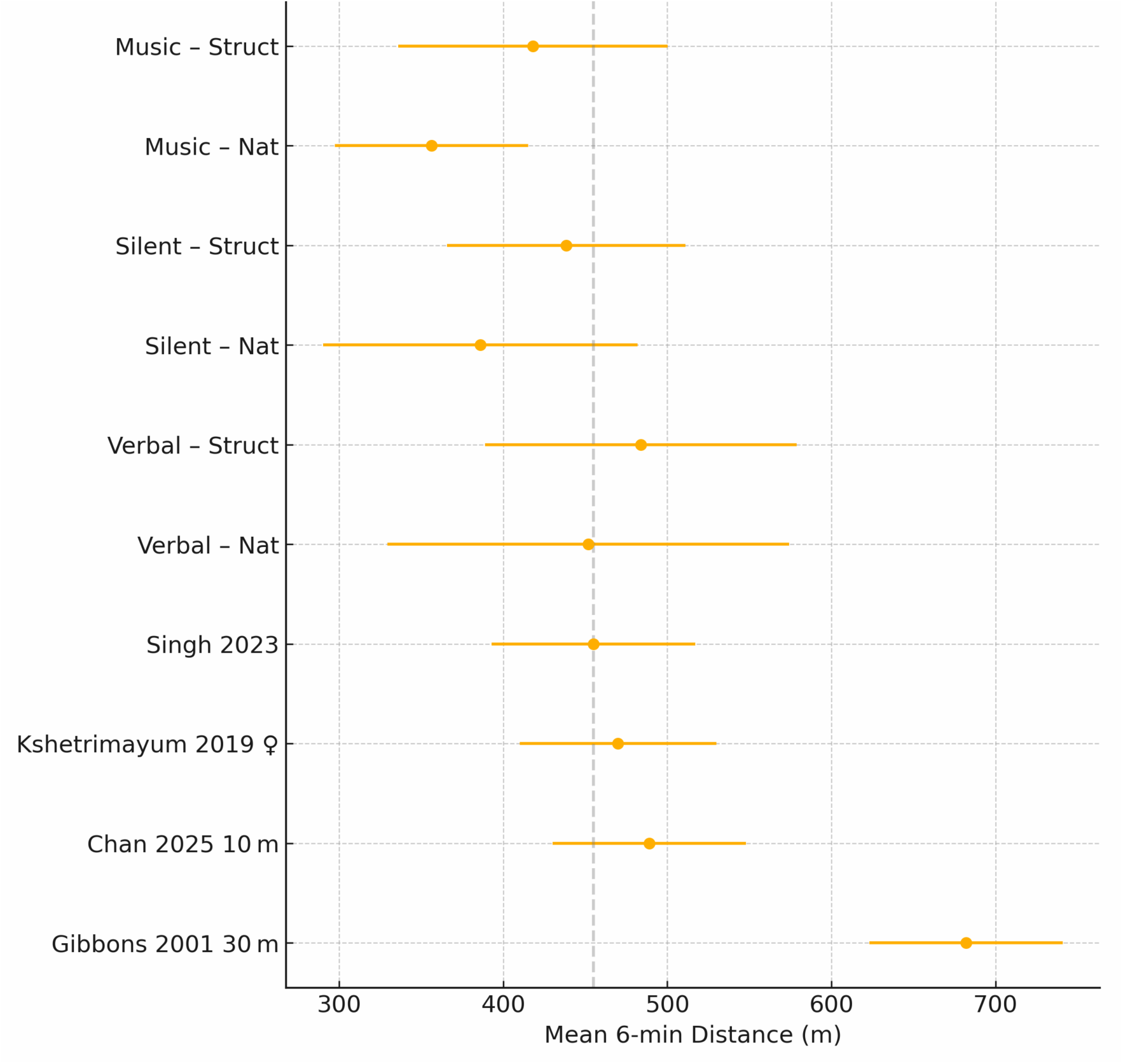
Forest plot – Ambulosono vs. published norms. Mean ± SD six minute walk distances for the six Ambulosono groups (Structured/Naturalistic × Silent/Verbal/Music) plotted alongside four reference values from the literature (Singh et al., 2023; Kshetrimayum & Saikia, 2019; Chan et al., 2025; Gibbons et al., 2001), illustrating alignment with short lane norms and divergence from classic 30 m corridor benchmarks.

### Sensitivity analyses

Repeating the environment comparison with raw, un-scaled distances (i.e., before six-minute normalisation) inflated structured means by 5–7 % yet still failed to cross the 0.05 significance threshold. Removing the nine longest naturalistic lanes (> 18 m) or the eight shortest structured lanes (< 10 m) did not change any p-value by more than 0.01, indicating that lane-length heterogeneity had little influence once the walk was time-scaled.

## Discussion

This study confirmed that the Ambulosono thigh mounted inertial sensor system delivers an environment agnostic six minute walk distances (6MWD) in healthy young adults. Our database consisted of 63 supervised (“Structured”) and 49 unsupervised (“Naturalistic”) walks from 76 participants. Mean scaled distances ranged from 388 m (Naturalistic–Silent) to 481 m (Structured–Verbal). Independent Welch t tests showed no significant differences between settings for any auditory condition: +61 m in Silent (t = 1.70, p = 0.10, d = 0.70), +52 m in Verbal (t = 1.68, p = 0.10, d = 0.39), and +37 m in Music (t = 1.04, p = 0.31, d = 0.43). All deltas lay within the device’s validated ± 5 % error margin (Hu, 2019; Chomiak et al., 2019) and below the 25–30 m minimal clinically important difference for young adults (Moura et al., 2019; Swigris et al., 2014).

### Influence of auditory cues

We also examined whether Silence, ATS style verbal encouragement, or rhythmic music differentially affected performance. Although Structured trials were consistently longer than Naturalistic trials across all conditions, the rank order of group means (Verbal > Music ≈ Silent) was identical in both environments (Figure 2). Verbal prompts and time reminders produced the highest average distances, while Music and Silent conditions were comparable. Crucially, there was no environment × auditory interaction—auditory stimuli influenced absolute pacing but did not alter the equivalence between settings. This finding implies that clinicians and researchers can choose any of these auditory paradigms without needing setting specific calibration.

### Agreement with the broader literature

Absolute 6MWD in our cohort (388–481 m) aligns with recent short lane norms for Asian young adults—455 ± 62 m on 20 m courses (Singh et al., 2023), 470 m in females (Kshetrimayum & Saikia, 2019), and 489 ± 59 m on a 10 m lane (Chan, Lee, & Mak, 2025)—yet remains 150–300 m below classic Western 30 m corridor norms (682 ± 59 m; Gibbons et al., 2001). A forest plot (Figure 6) highlights that lane geometry and anthropometry, rather than sensor under estimation, explain this discrepancy.

### Wearable sensors versus smartphones

Our environment independent results contrast sharply with smartphone only solutions. iPhone native sensor and apps underestimate 6MWD by ∼ 8–11 % in healthy and clinical cohorts (Ratliff et al., 2019; Zhang et al., 2023), and home based reliability drops to ICC = 0.74 (Mak et al., 2021). Apple’s own algorithm even caps estimates at 500 m (Apple, 2019; Apple Support, 2023). Ambulosono, by contrast, remained within ± 5 % error in both supervised and free living conditions, underscoring the value of dedicated inertial wearables for accurate digital health monitoring.

### Normative and clinical context

Across the lifespan and in disease, 6MWD varies widely. School aged children walk ∼ 470 m (Cacau et al., 2016); adolescents ∼ 550 m (Saraff et al., 2014); older adults 400–550 m (Voigt Radloff et al., 2016; Herdman et al., 2015). Clinical populations often fall below 400 m (McGavin et al., 1976; Morgan et al., 2010; Gijbels et al., 2010; Kägi et al., 2021; Takken et al., 2018). Our healthy young adult means occupy the upper tiers of these bands, reaffirming that Ambulosono captures true functional capacity rather than any artificial ceiling.

### Strengths and limitations

A key strength is the systematic manipulation of both environment and auditory cues, combined with rigorous time normalisation and benchmarking against lane matched norms. However, participant identifiers did not align across settings, precluding within subject Bland–Altman analysis.

Coefficient of step regularity was captured in fewer than half of trials, limiting mechanistic insight. The cohort’s youth and healthfulness also constrain generalisability to older or clinical populations. Facility constraints required occasional use of an 80 m circuit in the Structured setting, although normalisation mitigated this variation.

### Practical implications and future directions

Ambulosono enables reliable, corridor free 6MWT implementation: a 10–20 m home route yields the same time scaled distance as a supervised 12 m lane. Clinicians should interpret remote 6MWD against short lane reference values and can select Silent, Verbal or Music paradigms interchangeably. Future work should standardise identifiers across settings, improve CSR data capture, and extend validation to older adults and patient groups.

## Conclusion

Time scaled six minute walk distance measured by the Ambulosono sensor is robust to environment and auditory input and aligns with contemporary short lane normative values for healthy young adults. In contrast to smartphone only solutions that exhibit greater error and hard coded ceilings, Ambulosono meets clinical accuracy requirements, supporting its deployment for remote functional capacity monitoring in digital health and rehabilitation context.

## Data Availability

All data produced in the present study are available upon reasonable request to the authors

## Acknowledgements

This research was supported by CIHR, Alberta Innovation and Ministry of Mental Health and University of Calgary Endowment Fund

